# A web survey to assess the use efficacy of personnel protective materials among allied health care workers during COVID-19 pandemic at North-East India

**DOI:** 10.1101/2020.06.08.20125708

**Authors:** Rahul P Kotian, Manna Debnath, Zosangliani, Brayal D’souza, Disha Faujdar

## Abstract

The rising pandemic is resulting in increased usage of personnel protective equipment in the hospital and community. The efficient and effective use of appropriate personal protective equipment will help assure its availability and healthcare provider safety. The purpose of this study was to assess the use efficacy of PPE among health care workers through a web based survey during the pandemic. the response rate of the survey was 66.75%. 35.2% gave a full rating on a point of 5 regarding the control measures taken by the hospital, 39% of respondents did not use the PPE, 90.6% used a surgical mask while 65.9% wore the disposable gloves and only 47.6% wore the goggles/face shield More than half the respondents did not wear the shoe-cover. 97.4% used the hand sanitizer and around 97% maintained hand hygiene practice.

## 1. Introduction

The World Health Organization (WHO) declared COVID-19, the disease caused by SARS-CoV-2, a pandemic. The SARS-CoV-2 virus is spread through inoculation of mucous membranes by droplets and aerosols containing the virus (1), as well as contact with droplet-contaminated fomites (2,3). The world health organisation WHO on April 2020 has issued guidance on Rational use of personal protective equipment for coronavirus disease (COVID-19) and considerations during severe shortages. The WHO has recommended that when dealing with patients on a suspected COVID-19 positive patient must wear an N95 or FFP2 mask. There is also a recommendation that a medical mask, gown, gloves, and eye protection (goggles or face shield) is sufficient. The WHO also recommends that other staff on a ward not providing direct care require no PPE (4).

A narrative review by Dr T.M.cook on Personal protective equipment during the COVID −19 pandemic –summarises that Personal protective equipment is only one part of a system to protect staff and other patients from COVID −19 transmission, appropriate use of PPE significantly reduces risk of viral transmission and infection, Overuse of PPE is a form of misuse., misuse of PPE depletes limited stocks, leads to avoidable shortages and increases risk to staff (5). Shannon et al proposed p three types of PPE in their findings i.e. PPE for droplet and contact precautions, for general airborne, droplet, and contact precautions, and for those performing or assisting with high-risk aerosol-generating medical procedures (6). A Cochrane review on Protective clothes and equipment for healthcare workers to prevent them catching coronavirus and other highly infectious diseases resulted hat there very low-certainty evidence that covering more parts of the body leads to better protection resultant of difficult donning and duffing .Face-to-face training in PPE use may reduce errors more than folder-based training (7,8). A hospital based cross sectional study in n Oyo State, Nigeria. resulted that use of personal protective equipment (PPE) among their health care workers was low (56.8%) (9). A study in Uganda resulted in good practices among health care workers toward COVID-19 and age was a factor associated with good practise (10).

## 2. Methods

### 2.1 Participants

A web-based survey was conducted from 3^rd^ May to 10^th^ May during the lockdown period of North East India. During the lockdown period community based national sampling is not achievable so we decided to accumulate all the data online. By using the google form we prepared a set a questionnaire and send the link of the questionnaires to various online social media platform for collecting the data. Before circulating the google form a clear instruction was given to the participants about the purpose of the study. Participants had to answer yes or no in the inform consent before the formal survey, to confirm the willingness to participate voluntarily. After taking the consent the participants were directed to complete the self-survey questionnaire.

### 2.2 Ethical Considerations

Confidentiality of the study participants’ information was maintained throughout the study by making the participants’ information anonymous. An informed consent was obtained from each participant prior to participation. Participants were allowed to terminate the survey at any time they desired. The survey was anonymous, and confidentiality of information was assured.

### 2.3 Data Collection

The data collection was done between 3^rd^ May to 10^th^ May during the lockdown period of North East India. Clear instructions about details and purpose of the study was provided before the beginning of the questionnaires.

### 2.4 Measures

The questionnaire for the knowledge of control of COVID-19 and protective materials was developed by authors for north eastern allied health professionals. The questionnaires consisted of two parts: first part is demographic, and second part is knowledge based. The questionnaire had 8 questions and all the question are regarding controls and protective materials of COVID -19. Almost majority of the questionnaires were answer on Yes/No basis. Yes, means participants is having more knowledge of controls and protective materials and aware about corona outbreak. No means participants are having no adequate knowledge of controls and protective materials.

### 2.5 Statistical analysis

The analysis was done by using descriptive statistics. The analysis was performed by using Statistical Package for the Social Sciences SPSS (Version −20.0).

## 3. Results

A total of 400 Allied Healthcare Professionals in North-East India was requested to participate in the study through social media platforms. The questionnaire was circulated using Whatsapp and E-mails and around 267 responded. These respondents belonged to various professions including Medical Imaging, Physiotherapy, Emergency and Critical Care, Medical Lab Technology, Optometry, Operation Theatre Technology, and Nurses. Majority of the respondents were from medical imaging (68.2%). Among this total population, 55.4% were male while 44.6% were females.

**Table.1.**
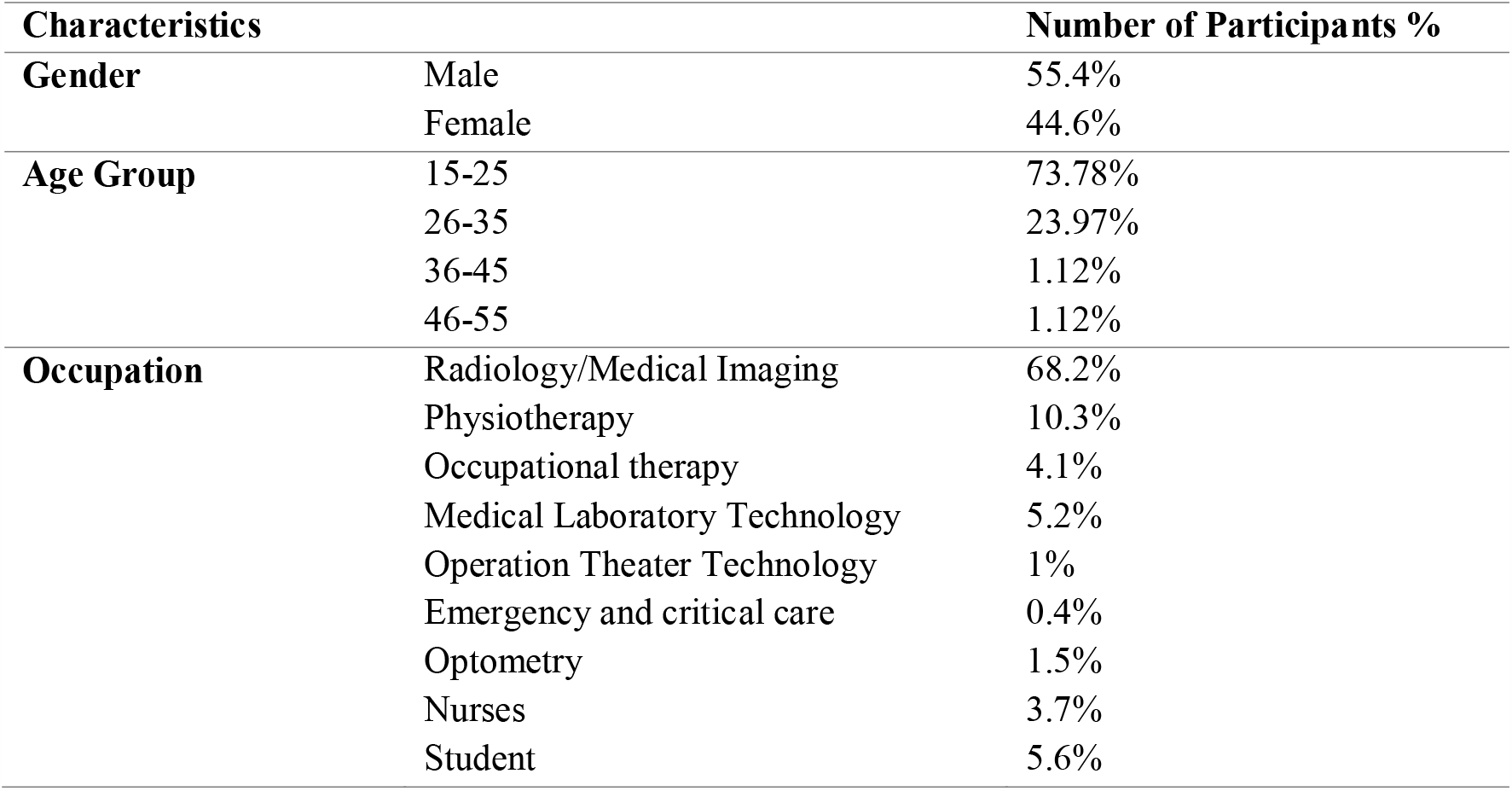
Demographic characteristic of participants.

Participants were asked to rate the control measures taken by their respective hospitals/ clinics/ center where, only 35.2% gave a full rating on a point of 5, 30.3% gave 4 out of 5, 20.6% awarded 3 out of 5, 6.7% gave 2and 7.1% awarded out of 5.

Around 95.9% respondents agreed that they were not in any direct contact with a COVID-19 patient. While 39% respondents answered that they did not wear the protective clothing. Most of the participants i.e. 90.6% agreed that they wear disposable surgical mask. While 65.9% wore the disposable gloves and only 47.6% wore the goggles/ face mask. More than half the respondents did not wear the shoe-cover. Almost all the respondents i.e. 97.4% were using the hand sanitizer and around 97% were maintaining hand hygiene.

**Table. 2.** Responses of Allied Health Professionals.

| Q.no | Questions | Options (%) |
| --- | --- | --- |
| **1** | Rate the amount of control measures taken for COVID-19 in your hospital/diagnostic center/clinics. | 1(7.1%), 2(6.7%), 3(20.6%), 4(30.3%), 5(35.2%) |
| **2** | Any direct contact with COVID-19 patient? | Yes (4.1%), No (95.9%) |
| **3** | Do you wear protective clothing/Isolation gown? | Yes (61%), No (39%) |
| **4** | Do you wear disposable surgical masks? | Yes (90.6%), No (9.4%) |
| **5** | Do you wear disposable latex gloves? | Yes (65.9%), No (34.1%) |
| **6** | Do you wear any protective goggle/face shield? | Yes (47.6%), No (52.4%) |
| **7** | Do you wear shoe cover? | Yes (46.4%), No (53.6%) |
| **8** | Do you maintain hand-hygiene? | Yes (97%), No (3%) |
| **9** | Do you use hand sanitizer? | Yes (97.4%), No (2.6%) |

## 4. Discussion

The coronavirus disease emerged in Wuhan, China and this disease is rapidly spreading from human to human. Our northeastern healthcare professionals are directly and indirectly working under this pandemic situation, so the control measures are very important among all the healthcare professionals and their respective hospitals and clinics for the prevention or the elimination of this deadly virus.

Kotian RP et al, performed a study among all the medical imaging professionals in which 45.6% workers underwent training session for the use of PPE, 91.5% professionals were well aware about all the important steps to prevent the transmission of COVID 19 (11). Zhong BL et al, in March 2020 conducted a study based on knowledge, attitude and practice towards COVID 19 outbreak in which they reported that the majority (98.0%) of the crowd are wearing mark in these circumstances and only few (2.0%) had not worn mark (12). Similarly, Roy D et al, 2020 performed a study based on COVID 19 pandemic they reported that 97% participants agreed that frequent hand wash can stop the prevent of infection. Furthermore 37% of participants informed that they are using mask without any early sign and symptoms of the disease, more than 75% participants felt that there is a need to use sanitizers and gloves and 85% agreed that they frequently wash their hands (13). In present study, majority of the participants i.e. 90.6% agreed that they wear disposable surgical mask, 65.9% participants wore disposable gloves and 47.6% wore face mask. More than half of the allied health workers from northeastern state are aware about wearing personal protective equipment’s in their workplace.

Bhagavathula AS et al, 2020 based on healthcare workers on which more than 85% of healthcare workers reported that hand hygiene, covering the nose and mouth while coughing can prevent healthcare transmission (14). Similarly, in present study we found that about 97.4% were using hand sanitizer and almost 97% participants were agreed that they were properly maintaining hand hygiene. This indicates that almost all the participants are well aware about the hand hygiene during this pandemic situation and concern about personal hygiene to avoid the COVID19 infection. The majority of participant’s i.e. 35.2% gave 5 out of 5 rating for the control measures taken by hospitals in north east India in terms of using all the PPE and sanitization of hospital premises and maintain social distance among all the medical staffs to avoid the COVID 19 infection.

Zhou M et al, April 2020, conducted a study among healthcare workers on which 89% of healthcare workers had the sufficient knowledge and 89.7% of healthcare workers followed all correct practices regarding COVID 19 (15). Shi Y et al, March 2020 89.51 % staff have good knowledge of COVID 19 and 64.63% of them received the COVID 19 training in their hospital (16). According to Giao H et al, 88.4% staff have good knowledge about COVID 19(17). In present study all the participants having more than average knowledge and control measures taken by their respective hospitals but 39% of participants responded that they did not wear the protective clothing and approximately 50% participants did not wear shoe cover. This study shows drastic control measures implemented by northeastern hospital to prevent the spread and reduce the risk of infection among all the healthcare professionals. To optimize the use of PPE there should be an increase in adherence to protocols for PPE use, improved PPE design, and research into the risks, benefits, and best practices of PPE use (18).

A rapid systematic review on the efficacy of face masks and respirators against coronaviruses and other respiratory transmissible viruses for the community, healthcare workers and sick patientsresulted that in healthcare settings continuous use of respirators, is more protective compared to the medical masks, and medical masks are more protective than cloth masks. Cloth masks may not be safe for health workers as it depends on the fabric and design (19).

## 5. Conclusion

The study assessed the use efficacy among health care workers, it is also important to study the errors made in removal and discard that can increase the risk of contamination and any contributing physical discomfort that prevents or minimizes the use of PPE among health care workers

## Data Availability

The raw data was interpreted during the present study is made available by the corresponding author and uploaded in the supplementary files.

## Acknowledgements

The authors are thankful to all the allied healthcare professionals from the North-Eastern India for actively participating in the survey study.

The authors thank all the Healthcare professionals involved in this study for their cooperation and support.

## ‘Declarations’

### Ethics approval and consent to participate

The study protocol followed was reviewed and approved by the Research Committee of Srinivas University telephonically and approved online. The consent to participate approval was also taken.

### Ehical approval reference number

Not applicable

### Consent for publication

A detailed explanation about the study was given by the principal investigator after which they provided consent for publication. All the patients included in this research gave written informed consent to publish the data contained within this study.

### Availability of data and material

The datasets used and/or analysed during the current study is made available by the corresponding author and attached in the supplementary files.

### Authors’ contributions

RK conceptualized the study. DF, MD and Z have given inputs in study design. Z, MD, and DF collected the data. RK analysed the data and wrote the first draft of manuscript and all co-authors contributed in critical review of data analysis and manuscript writing. RK will act as guarantor for this paper.

